# Mortality in cancer patients with congenital anomalies across different age groups: trend analysis and prognostic risk factors

**DOI:** 10.1101/2021.11.20.21266629

**Authors:** Sara Morsy, Truong Hong Hieu, Sherief Ghozy, Linh Tran, Nguyen Tien Huy

**Affiliations:** Medical Biochemistry and Molecular Biology Department, Faculty of Medicine, Tanta University, Egypt; Faculty of Medicine, University of Medicine and Pharmacy at Ho Chi Minh City: Ho Chi Minh, Vietnam; Neurosurgery Department, El Sheikh Zayed Specialized Hospital, Giza, Egypt; Institute of Research and Development, Duy Tan University, Da Nang, Vietnam; Evidence Based Medicine Research Group, Ton Duc Thang University, Ho Chi Minh City, 70000, Vietnam; Faculty of Applied Sciences, Ton Duc Thang University, Ho Chi Minh City, 70000, Vietnam; Department of Clinical Product Development, Institute of Tropical Medicine (NEKKEN), School of Tropical Medicine and Global Health, Nagasaki University, Nagasaki 852-8523, Japan

**Keywords:** cancers, congenital anomalies, death, prognostic factors, SEER

## Abstract

**Purpose:** Congenital anomalies are one of the causes of the high mortality rate in children diagnosed with cancer. However, there is a gap of evidence of the rate of cancer mortality in older patients who had congenital anomalies. The study, therefore, aimed to investigate the epidemiology of cancer mortality in those patients.

**Methods:** Data were retrieved for patients with cancer and died due to congenital causes throughout 43 years from Surveillance, Epidemiology, and End Results program SEER. The age of patients was divided into nine groups each is formed of 10 years interval. Joinpoint analysis was used to calculate the trends of Cancer mortality and Cox proportional hazard ratio to identify the mortality risk factors.

**Results:** We have included 2682 patients with death associated with congenital malformation. The mortality of cancer patients due to congenital anomalies greatly enhanced in the last years with the overall average annual percent was 3.8%. Interestingly, congenital anomalies had less mortality risk than other causes reported in SEER. Moreover, age, sex, radiation, chemotherapy, and behavior of tumor were significantly associated with higher survival in patients with congenital anomalies.

**Conclusions:** Cancer patients with congenital anomalies had less mortality risk than patients with other diseases reported in SEER. The mortality rates decreased recently, with the most mortality in the bone marrow and prostate tumors.

**Implications for Cancer Survivors:** Congenital anomalies are considered the least studied diseases in cancer patients. In this study, we studied how congenital anomalies did not increase the risk for cancer. However, our analysis implied the congenital anomalies in the male reproductive system were associated with the highest risk of cancer.

## Introduction

Congenital anomalies, which are diagnosed in at least 120,000 infants annually in the United States, are a major cause of infant death as well as pediatric hospitalizations [1-3]. The maternal diagnosis of cancer, whether before or after pregnancy, has been associated with a higher risk of congenital anomalies in the offspring [4]. Although the etiology of childhood malignancies remains poorly understood, several studies have provided evidence for an increased risk of cancer in children with congenital anomalies [5, 6]. The aforementioned association may suggest potential cancer-predisposing conditions and further involvement of developmental genetic mechanisms [7, 8]. There are several different ways in which the presence of birth defects might lead to the risk of childhood cancer development, including shared genetic and/or environmental factors, or diversity in organ structure or function, or through adapted lifestyle related to human disability. For instance, homeobox genes were among the genes implicated in congenital anomalies and also were related to a high risk of cancer [9]. It has been also found that patients with chromosomal aberrations had a higher risk for leukemia while non-chromosomal abnormalities had a higher risk for non-Hodgkin lymphoma [10].

Several congenital anomalies have been associated with certain cancers risk. For example, Down syndrome is associated with the risk of specific leukemias, autosomal deletion with retinoblastoma, and Beckwith-Weideman syndrome with Wilms’ tumor [11, 12]. Moreover, various published studies have reported that patients with congenital anomalies have an increased risk of developing cancer, such as leukemia, lymphoma, brain tumor, neuroblastoma, germ cell tumor, retinoblastoma, and soft tissue sarcoma [2, 13-18]. The risk of congenital anomalies has been suggested to be higher in infants born to mothers who were diagnosed with pregnancy-associated cancer [19], including lymphomas [20] and breast cancer [21]; however, the results have not been consistent and more studies are needed [22, 4]. A study found that there is a decreased risk of tumors due to congenital anomalies in adolescents due to less effect of developmental genes in this age group [10].

Despite the increasing evidence that the presence of congenital anomalies increased the mortality in childhood tumors, there is little or no evidence indicating the mortality risk in older patients. Therefore, this study aims to investigate the trend and prognostic risk of cancer mortality in patients with congenital anomalies. We also would investigate the age groups associated with higher mortality and whether the high mortality rate is only limited to a younger age.

## Methods

### Study population and characteristics

Data were extracted from SEER database using SEER*stat 8.3.6. The extracted data included marital status, race, age, chemotherapy, radiation, sequence, surgery, site, laterality, pathology, sex, and behavior regarding the cancer patients who died due to congenital anomalies. For the trend analysis, the year of death due to congenital anomalies was calculated as the sum of the year of cancer diagnosis and survival months. We extracted the age of the patients and divided it into nine intervals of 10 years each.

For comparison to the other causes of death in SEER database, we matched the cases with our cohort of congenital anomalies for age and sex with rate of one to one so each cohort had 2682 cases using MatchIt package in R [23].

### Statistical analysis

All analyses were conducted using R 3.6.0 [24]. The categorical variables were expressed in percentage and continuous variables in mean and standard deviation (SD); otherwise, median and interquartile range were used. Trend analysis was used to calculate the annual cancer deaths due to congenital anomalies. First, we extracted the diagnosis year and survival months then we calculated the year of death for each patient. Moreover, trend analysis was conducted using joinpoint regression analysis with four join points and annual percentage change (APC) was calculated and its 95% confidence interval [25]. The joinpoint regression analysis was conducted using Joinpoint 4.6.0 [26].

Missing data were imputed using the KNN algorithm with several neighbors equal to 5. The imputation was conducted in R using VIM library [27]. In addition, a Proportional Hazards Cox Regression analysis was performed, and the accuracy of the model was tested using the concordance index. The significant variables resulting from univariate Cox regression analysis were used to construct multivariate Cox regression.

A survival decision tree was constructed using rpart package [28]; we used the minimum variable at each split of 10 and the maximum depth of 10 then we pruned the tree to avoid overfitting. The prediction error was calculated using an integrated brier score using ipred package [29]. Results were considered significant when the *p-value* was less than 0.05.

## Results

We identified 2682 cases that died due to congenital anomalies from 1975 to 2018. The cohort included 1511 males representing 56.3% of the total cohort Table 1. The mean age of cancer patients who died due to congenital anomalies was 60.6 years old. We grouped the patients based on age into nine groups; each group has 10 years interval. Interestingly, we found that most cancer patients who died due to congenital anomalies were old patients (range: 70 – 80 years old). We found that the age group from 10 to 20 years old were the least group to die due to congenital anomalies. Most patients were married (52.9), white (68.2), and had a malignant tumor (90.2%). Most patients received radiotherapy and surgery but not chemotherapy Table 1.

**Table 1.**
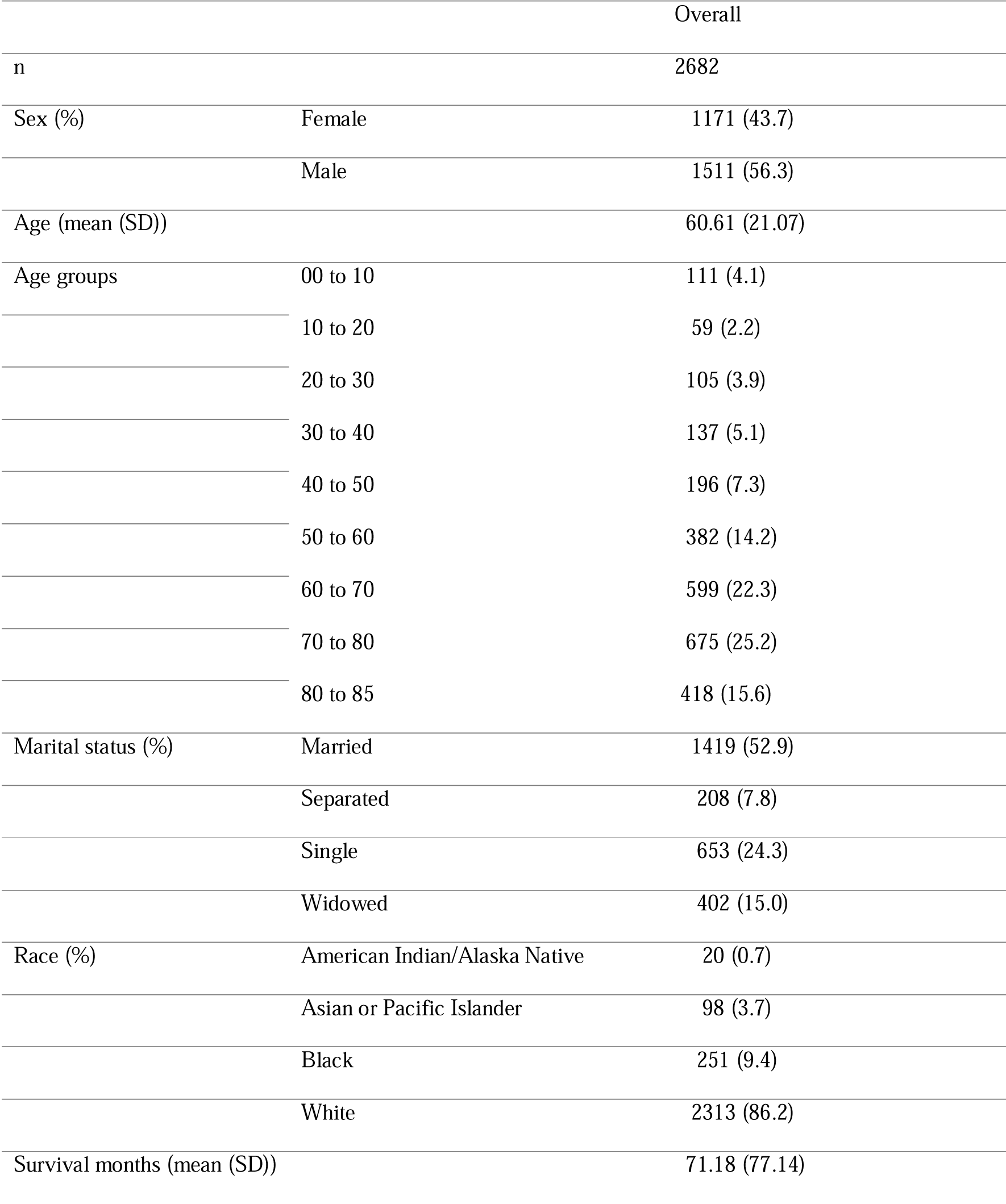
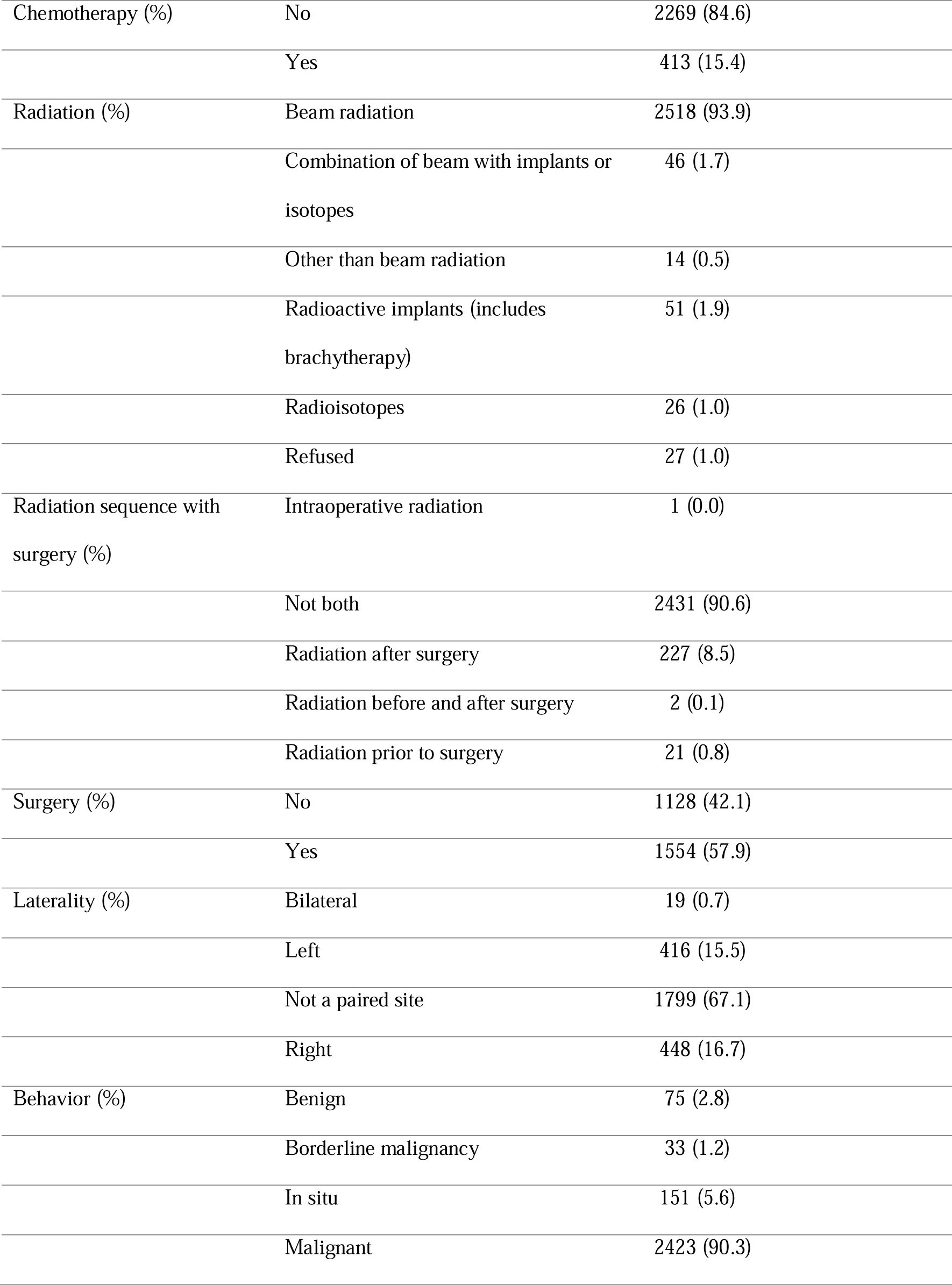
Characteristics of included patients.

Furthermore, adenoma and adenocarcinoma had the highest numbers among patients who died due to congenital anomalies (n = 909) (Figure 1).

**Figure 1.**
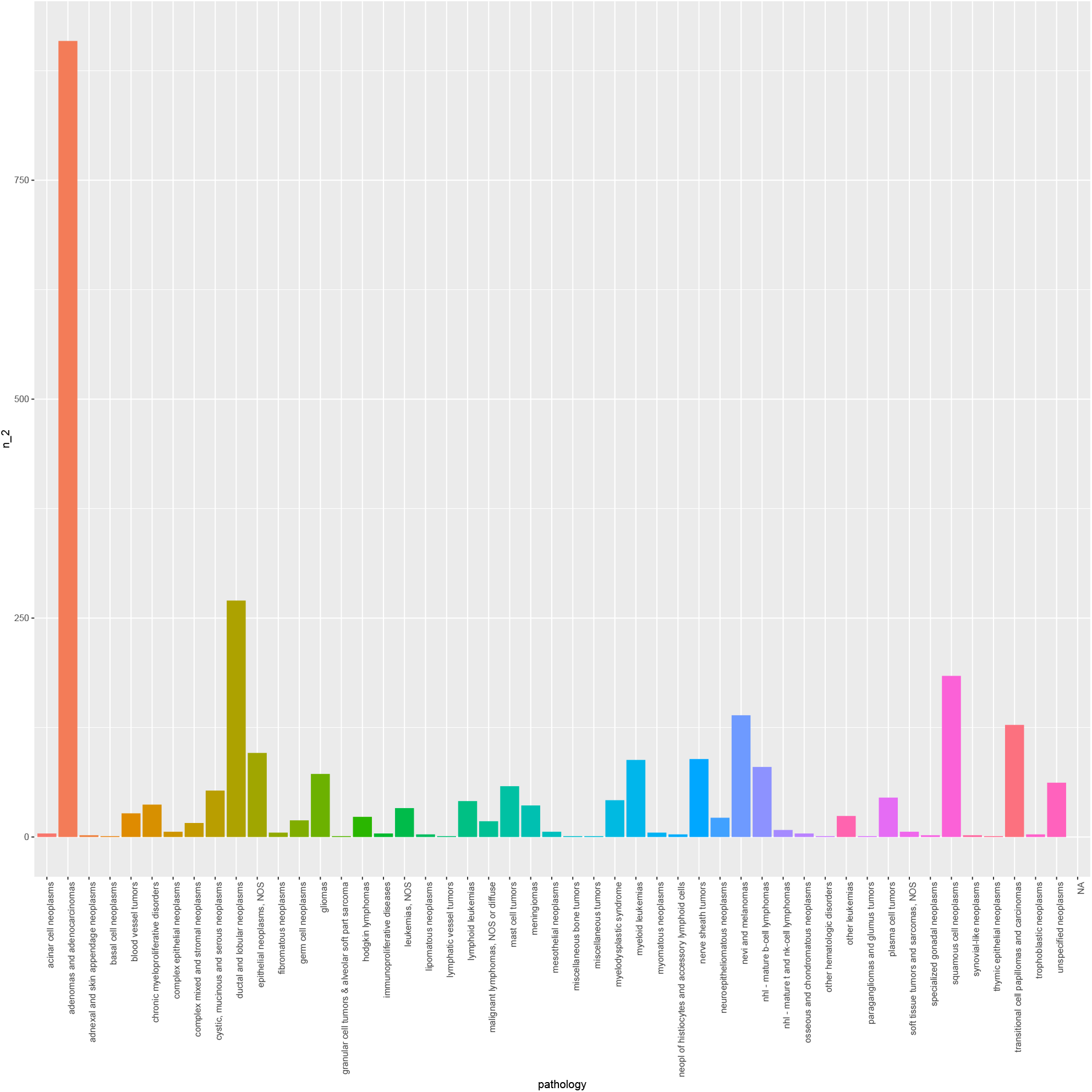
The number of mortality cases due to congenital anomalies based on the pathology of tumor

The male genital system was the site for most of the adenoma and adenocarcinoma cases representing 34% of all cases. Meanwhile, bone marrow (n = 362) and prostate (n = 337) were the most common site associated with mortality (Supplementary figure 1A, B).

### Trends and incidence of cancer deaths due to congenital anomalies

The standardized mortality rate was 1.16 [95% CI (0.1; 1.33)]. The annual percent increase of mortality cases was 14.81% [95% CI (11.6; 18.1), *p-value < 0*.*001*] from 1975 to 1993 which was significant increase that continues till 2015 [4.3% [95% CI (2; 6.7), *p-value < 0*.*001*] (Figure 2). However, massive significant decline was observed from 2015 to 2017 [-60.4, (95% CI = -83.7; -4.1), *p-value <0*.*001*] (Figure 2).

**Figure 2.**
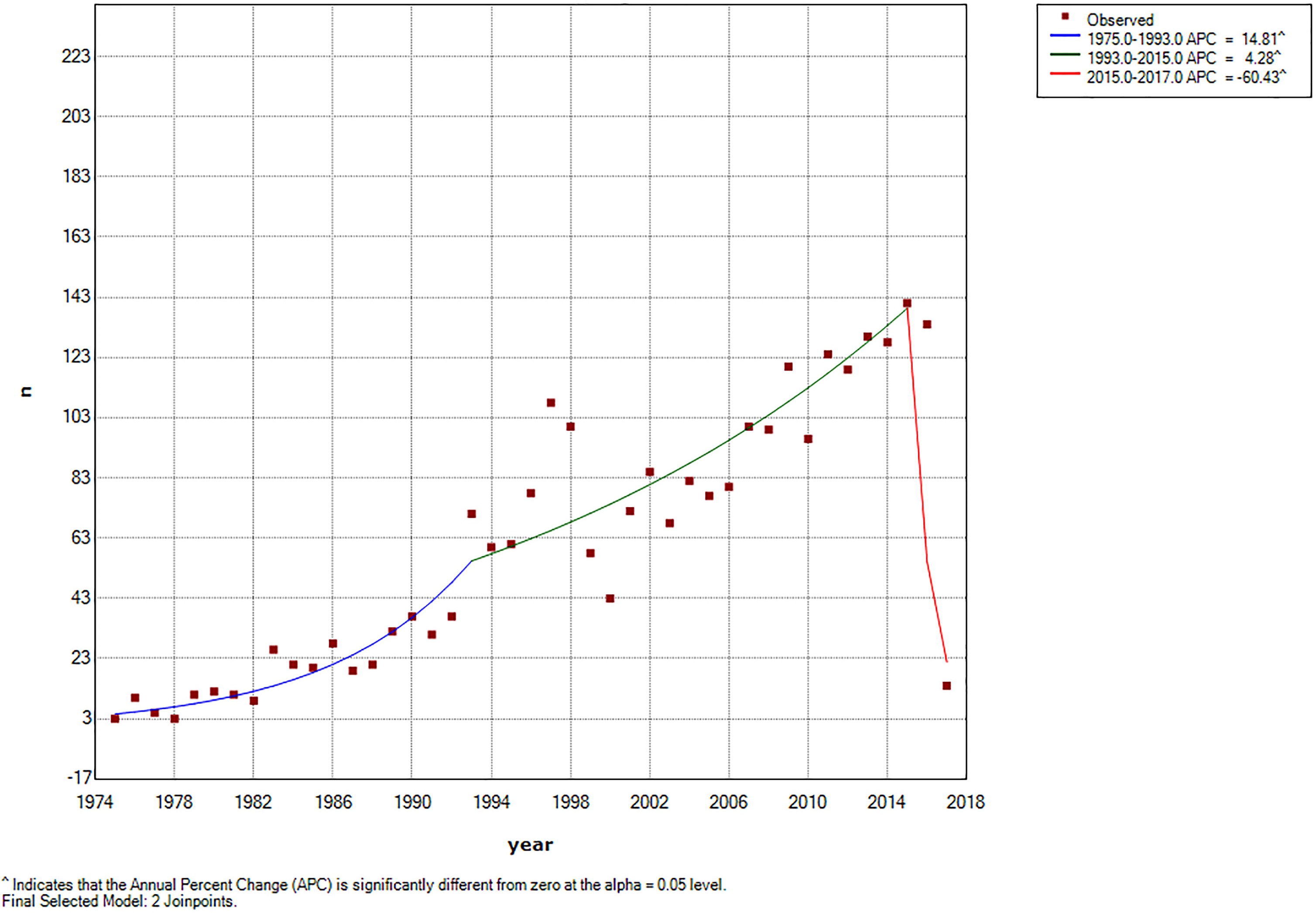
Joinpoint regression trend analysis of congenital anomalies caused deaths in cancer

### Comparison between mortality due to congenital anomalies and other causes

We matched a cohort of cases that died due to causes other than congenital anomalies. Our analysis revealed that there was a significant difference between survival duration for both cases (71 months for congenital analysis versus 92.52 for other causes, *p < 0*.*001*). Malignant tumors were significantly less prevalent in congenital anomalies Table 2. Other significant variables are present in Table 2.

**Table 2.**
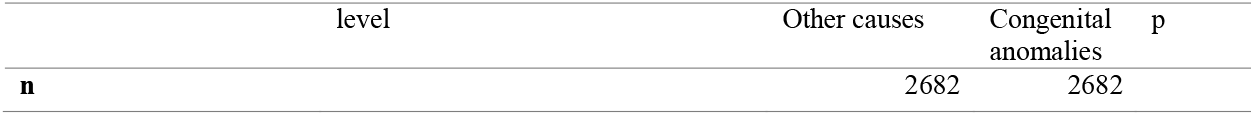
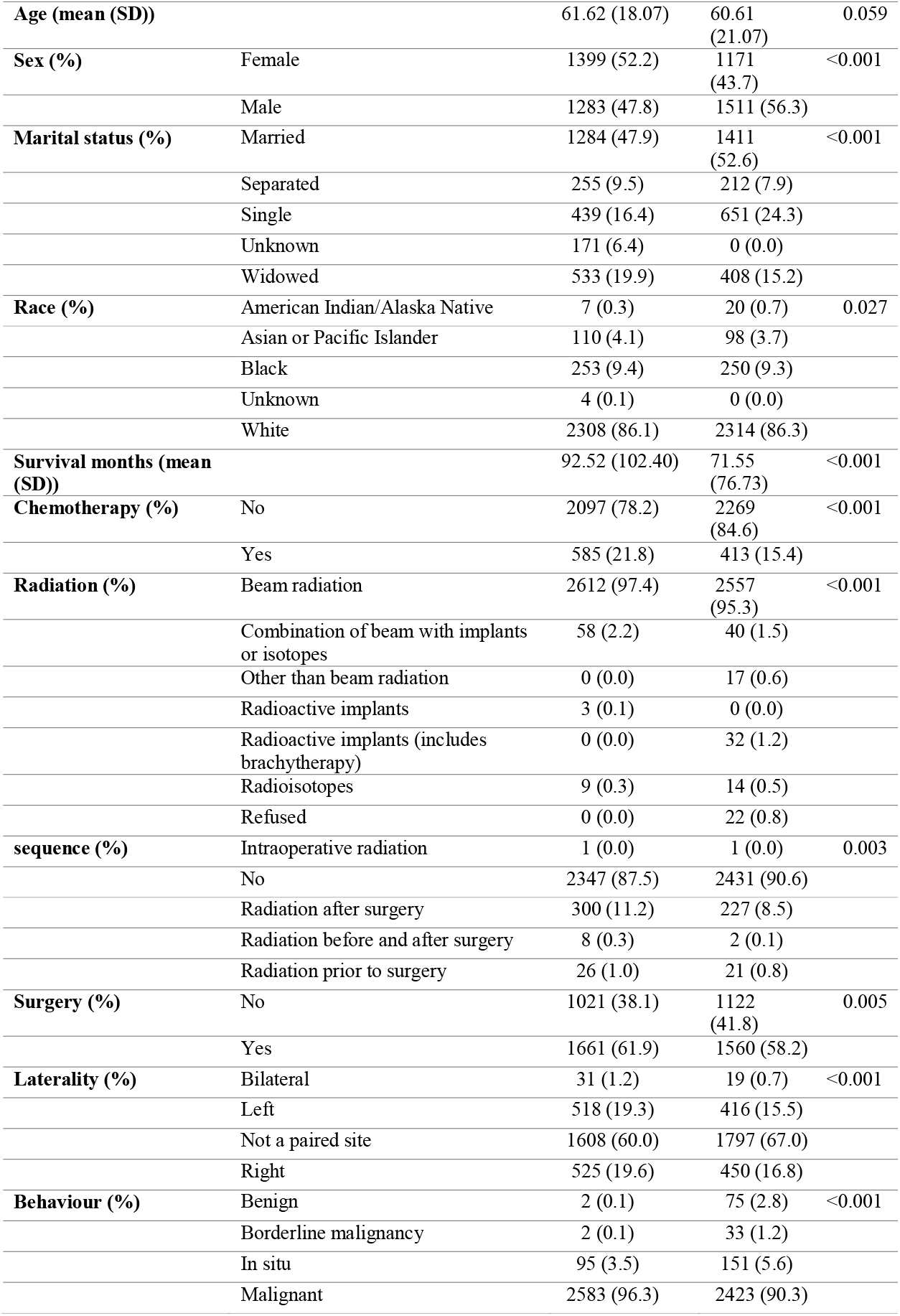
The difference between cancer mortality due to congenital anomalies or other causes.

Comparing the survival curves for cancer patients who died due to congenital anomalies and others who died due to other causes (in a matched cohort), congenital anomalies had less mortality risk than other causes reported in SEER database [HR = 1.28, SE = 0.03, p < 0.001] (Figure 3).

**Figure 3.**
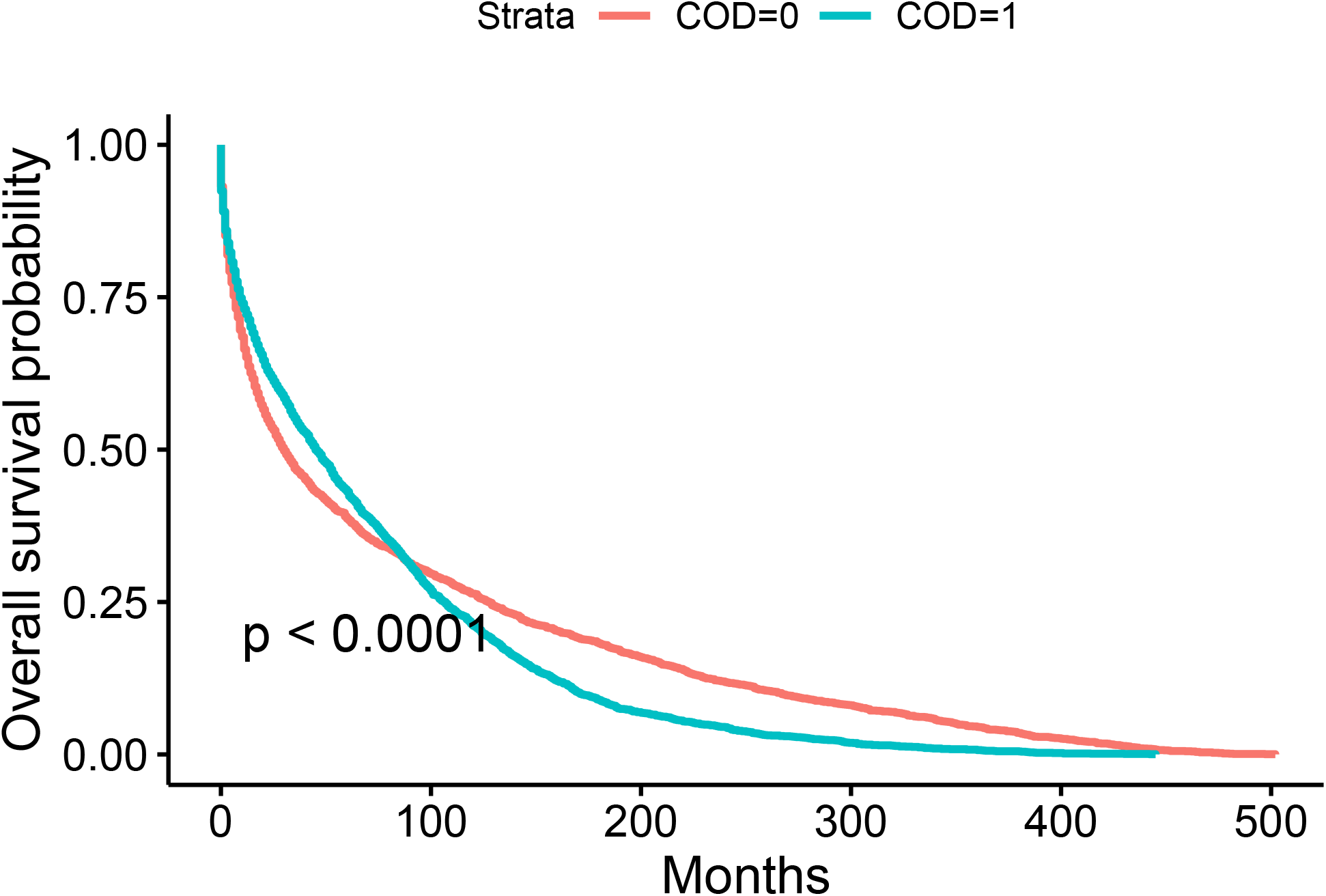
Kaplan Meier analysis showing that congenital anomalies (1) had less mortality risk than other causes (0).

However, comparing the survival curves between congenital anomalies and others who died due to other causes in specific cancers e.g. breast adenomas and GIT adenomas did not reveal any significant difference (Supplementary figure 2A, B).

Univariate cox regression analysis implied that only age, sex, radiation, marital status chemotherapy, and behavior of the tumor have affected the survival of these patients. Stratifying age into different groups implied that the highest mortality risk group was from > 80 years old [HR = 1.34, SE = 0.12, p = 0.01] (Figure 4). Other groups had less risk of mortality than the first decade of life Table 3. In addition, these variables maintained their importance in the multivariate cox regression analysis.

**Figure 4.**
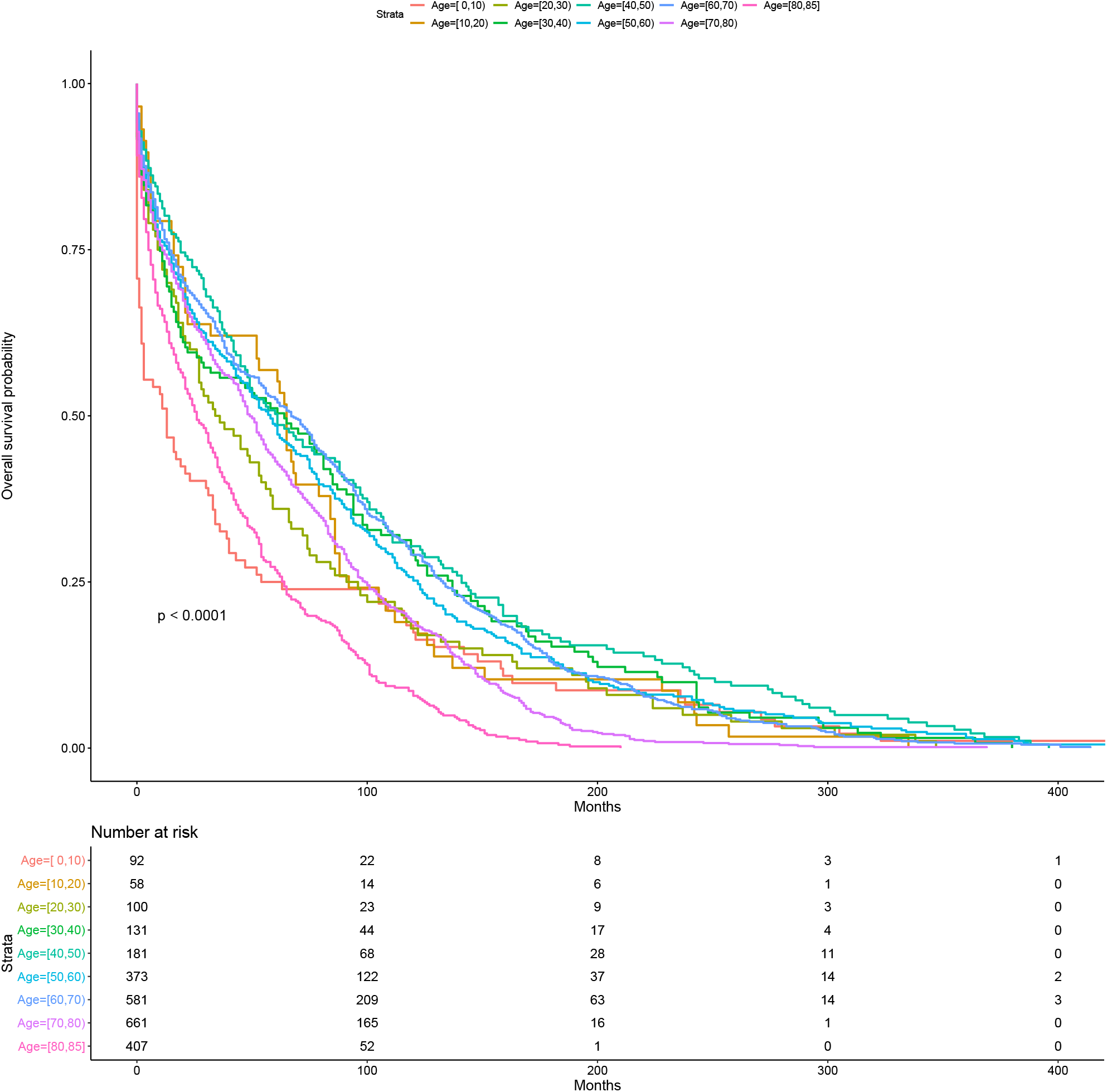
Kaplan Meier analysis showing the survival probability for each age group

**Table 3.**
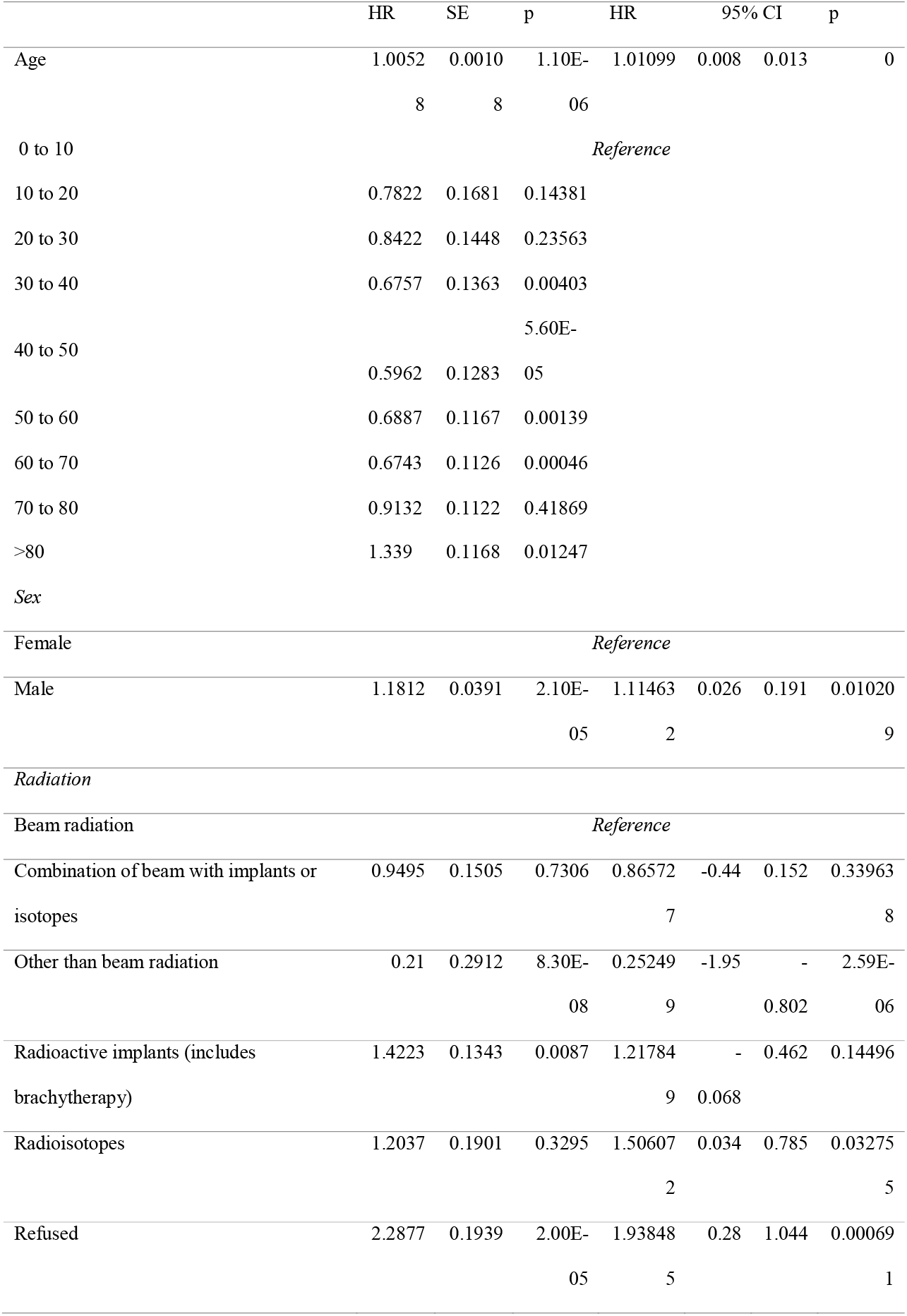
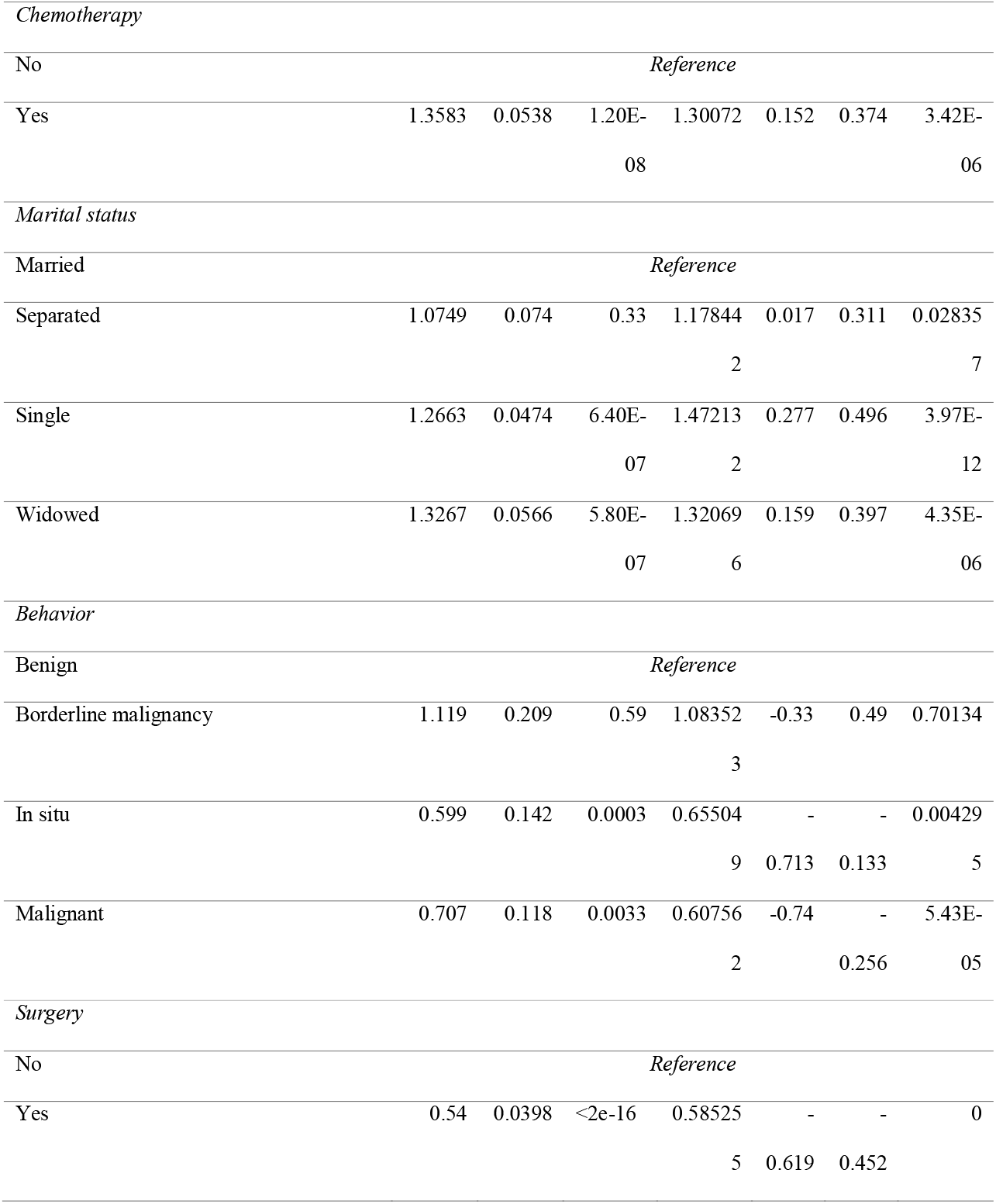
Univariable and multivariable cox regression analysis.

## Discussion

Our study has revealed that most patients with congenital anomalies have adenoma or adenocarcinoma. Higher mortality rates were observed in Bone marrow and prostate cancer. There were several factors, including age, sex, radiation, chemotherapy, and behavior of tumor having an association with the survival of cancer patients. In addition, the trend analysis revealed that there was a decline in cancer mortality caused by congenital anomalies in recent years. We could not find an explanation for this decline in the recent year from information present in the SEER database.

According to Dolk. H et.al. reported 2.5% of live births with congenital anomaly died in the first week of life and 2.0% were stillbirths or fetal deaths from the second trimester and terminations of pregnancy following prenatal diagnosis occurred approximately 20% of all patients [30]. Especially, the live neonates accounted for an overwhelming percentage with 80% [31]. Followed that, despite the fact is that having birth defects was associated with a rise in the risk of childhood cancer [5, 6], in our study, an intriguing finding pointed out throughout ten years of research there are only 2682 cancer patients died related to congenital anomalies and a group of age from 70-year-old to 80-year-old had the largest percentage, accounting for more than 25%. Meanwhile, only 4.1% and 2.2% of cancer patients aged 0 to 10 and 10 to 20, respectively, have the death attributed for this reason.

The previous study reported that the age of the patient is essential for evaluating cumulation and/or exposure to carcinogenic factors combined with the presence of congenital anomalies [32].

The results of our study supported that there was a worse survival in unmarried cancer patients who died because of congenital anomalies, which is supported by the result of the previous study [36]. This might result from less social support, leading to decrease treatment adherence. Furthermore, lack of social support also affected immune and endocrine function which played a critical role in the growth and progression of cancer [37].

The data analysis of our study showed chemotherapy and radiation have increased the risk of mortality. Since patients received these therapies were often at the late stage or unresectable or metastatic or contraindications of curative treatment as surgery. Another reason worth mentioning is that the variable side effects of chemotherapy can increase mortality rate [38].

Besides, although during 42 years from 1975 to 2017, the annual percent of mortality cases increase by less than 4%, in two years from 2015 to 2017, the number dropped significantly [-60.4, (95% CI = -83.7; -4.1), *p-value <0*.*001*]. This might be a result of different trends of incidence and mortality of cancers.

This study figured out those patients between 80 and 85 years old who had birth defects were at the highest risk of death which was similar to the general cancer patients [39]. In addition, bone marrow is the most common site associated with mortality in those cancer patients with death-related congenital malformations. Interestingly, a study in New York State figured out acute myeloid leukemia (AML) children with down syndrome showed better survival in comparison to children with other birth defects or no birth defects [41]. The reasonable explanation is typically genetic and other biological features, having a superior response to chemotherapeutic agents in the treatment and relapse frequent seen in DS children was low [43, 44]. Thus, deaths caused by those birth defects are possibly attributed to the poor survival outcome observed for AML patients with other congenital anomalies.

Adenoma and adenocarcinoma had the highest proportion among patients who died due to congenital anomalies. The male genital system comprised most of the adenoma and adenocarcinoma cases representing 34% of cases. The possible cause of this is at birth, cryptorchidism is the most frequent male genital disorder presented at birth, affecting 0.1–9% of all male newborns [45]. It is also one of the few known risk factors for testicular germ cell tumors (TGCT). It has been postulated that other congenital malformations, particularly hypospadias, are also related to a higher risk of cancer [46-48]. Moreover, data showed that the risk of TGCT rises significantly in urogenital abnormalities, specifically cryptorchidism, hypospadias, and inguinal hernia patients [48, 49].

## Data Availability

The data are available and can be accessed through the SEER database, which is publicly available at https://seer.cancer.gov/data/.

https://seer.cancer.gov/

## Compliance with Ethical statement

Not applicable

## Informed consent

Not applicable

## Ethical approval

Not applicable

## Author Contributions

Study concept and design, acquisition of the data, statistical analysis: Sara Morsy

Review of analysis: Nguyen Tien Huy, Sherief Ghozy

Writing and interpretation of data: Truong Hieu, Linh Tran, Sara Morsy, Sherief Ghozy, Nguyen Tien Huy Study supervision, critical revision of the manuscript for important intellectual content: Nguyen Tien Huy

## Figure legends

**Supplementary figure 1 A, B**. Case mortality numbers due to congenital anomalies recorded in each site of cancer (Supplemental files)

**Supplementary figure 2**. Kaplan Meier curves comparing survival probability of congenital anomalies and other causes of death for GIT adenoma (A) and breast adenoma (B) (Supplemental files)

## References

1. Wong-Siegel JR, Johnson KJ, Gettinger K, Cousins N, McAmis N, Zamarione A et al. Congenital neurodevelopmental anomalies in pediatric and young adult cancer. Am J Med Genet A. 2017;173(10):2670–9. doi:10.1002/ajmg.a.38387.

2. Braga M, Gianotti L, Vignali A, Di Carlo V. Preoperative oral arginine and n-3 fatty acid supplementation improves the immunometabolic host response and outcome after colorectal resection for cancer. Surgery. 2002;132(5):805–14. doi:10.1067/msy.2002.128350.

3. Hoyert DL, Mathews TJ, Menacker F, Strobino DM, Guyer B. Annual Summary of Vital Statistics: 2004. Pediatrics. 2006;117(1):168. doi:10.1542/peds.2005-2587.

4. Momen NC, Ernst A, Arendt LH, Olsen J, Li J, Gissler M et al. Maternal cancer and congenital anomalies in children - a Danish nationwide cohort study. PLoS One. 2017;12(3):e0173355–e. doi:10.1371/journal.pone.0173355.

5. Bjørge T, Cnattingius S, Lie RT, Tretli S, Engeland A. Cancer Risk in Children with Birth Defects and in Their Families: A Population Based Cohort Study of 5.2 Million Children from Norway and Sweden. Cancer Epidemiology Biomarkers &amp;amp; Prevention. 2008;17(3):500. doi:10.1158/1055-9965.EPI-07-2630.

6. Carozza SE, Langlois PH, Miller EA, Canfield M. Are Children With Birth Defects at Higher Risk of Childhood Cancers? American Journal of Epidemiology. 2012;175(12):1217–24. doi:10.1093/aje/kwr470.

7. Chmielecki J, Bailey M, He J, Elvin J, Vergilio JA, Ramkissoon S et al. Genomic Profiling of a Large Set of Diverse Pediatric Cancers Identifies Known and Novel Mutations across Tumor Spectra. Cancer Res. 2017;77(2):509–19. doi:10.1158/0008-5472.CAN-16-1106.

8. Sun Y, Overvad K, Olsen J. Cancer risks in children with congenital malformations in the nervous and circulatory system-A population based cohort study. Cancer Epidemiol. 2014;38(4):393–400. doi:10.1016/j.canep.2014.04.001.

9. Anbazhagan R, Raman V. Homeobox genes: Molecular link between congenital anomalies and cancer. European Journal of Cancer. 1997;33(4):635–7. doi:https://doi.org/10.1016/S0959-8049(97)00010-5.

10. Von Behren J, Fisher PG, Carmichael SL, Shaw GM, Reynolds P. An Investigation of Connections between Birth Defects and Cancers Arising in Adolescence and Very Young Adulthood. The Journal of Pediatrics. 2017;185:237–40. doi:https://doi.org/10.1016/j.jpeds.2017.02.057.

11. Miller RW. Relation between Cancer and Congenital Defects in Man. New England Journal of Medicine. 1966;275(2):87–93. doi:10.1056/nejm196607142750208.

12. Mili F, Khoury MJ, Flanders WD, Greenberg RS. Risk of Childhood Cancer for Infants with Birth Defects: I. A Record–Linkage Study, Atlanta, Georgia, 1968–1988. American Journal of Epidemiology. 1993;137(6):629–38. doi:10.1093/oxfordjournals.aje.a116720.

13. Fisher PG, Reynolds P, Von Behren J, Carmichael SL, Rasmussen SA, Shaw GM. Cancer in children with nonchromosomal birth defects. J Pediatr. 2012;160(6):978–83. doi:10.1016/j.jpeds.2011.12.006.

14. Narod SA, Hawkins MM, Robertson CM, Stiller CA. Congenital anomalies and childhood cancer in Great Britain. Am J Hum Genet. 1997;60(3):474–85.

15. Altmann AE, Halliday JL, Giles GG. Associations between congenital malformations and childhood cancer. A register-based case-control study. Br J Cancer. 1998;78(9):1244–9. doi:10.1038/bjc.1998.662.

16. Agha MM, Williams JI, Marrett L, To T, Zipursky A, Dodds L. Congenital abnormalities and childhood cancer. Cancer. 2005;103(9):1939–48. doi:10.1002/cncr.20985.

17. Berbel-Tornero O, Ortega-Garcia JA, Ferris-Tortajada J. Congenital abnormalities and childhood cancer: a cohort record-linkage study. Cancer. 2006;106(6):1418-9; author reply 9. doi:10.1002/cncr.21685.

18. Rankin J, Silf KA, Pearce MS, Parker L, Ward Platt M. Congenital anomaly and childhood cancer: A population-based, record linkage study. Pediatr Blood Cancer. 2008;51(5):608–12. doi:10.1002/pbc.21682.

19. Cardonick E, Usmani A, Ghaffar S. Perinatal outcomes of a pregnancy complicated by cancer, including neonatal follow-up after in utero exposure to chemotherapy: results of an international registry. Am J Clin Oncol. 2010;33(3):221–8. doi:10.1097/COC.0b013e3181a44ca9.

20. Langagergaard V, Horvath-Puho E, Norgaard M, Norgard B, Sorensen HT. Hodgkin’s disease and birth outcome: a Danish nationwide cohort study. Br J Cancer. 2008;98(1):183–8. doi:10.1038/sj.bjc.6604126.

21. Dalberg K, Eriksson J, Holmberg L. Birth outcome in women with previously treated breast cancer--a population-based cohort study from Sweden. PLoS Med. 2006;3(9):e336. doi:10.1371/journal.pmed.0030336.

22. Langagergaard V, Gislum M, Skriver MV, Norgard B, Lash TL, Rothman KJ et al. Birth outcome in women with breast cancer. Br J Cancer. 2006;94(1):142–6. doi:10.1038/sj.bjc.6602878.

23. Ho D, Imai K, King G, Stuart EA. MatchIt: Nonparametric Preprocessing for Parametric Causal Inference. Journal of Statistical Software; Vol 1, Issue 8 (2011). 2011.

24. Team RC. R: A Language and Environment for Statistical Computing. 2019.

25. Kim HJ, Fay MP, Feuer EJ, Midthune DN. Permutation tests for joinpoint regression with applications to cancer rates. Statistics in medicine. 2000;19(3):335–51. doi:10.1002/(sici)1097-0258(20000215)19:3<335::aid-sim336>3.0.co;2-z.

26. Joinpoint Regression Program. 4.6.0.0 ed: Statistical research and Applications Branch, National Cancer Institute.; 2018.

27. Templ AKaM. Imputation with the {R} Package {VIM}. Journal of Statistical Software. 2016;74. doi:10.18637/jss.v074.i07.

28. <rpart.pdf>.

29. Hothorn APaT. ipred: Improved Predictors. 2017.

30. Dolk H, Loane M, Garne E. The prevalence of congenital anomalies in Europe. Advances in experimental medicine and biology. 2010;686:349–64. doi:10.1007/978-90-481-9485-8_20.

31. Dolk H, Loane M, Garne E. The prevalence of congenital anomalies in Europe. Adv Exp Med Biol. 2010;686:349–64. doi:10.1007/978-90-481-9485-8_20.

32. Franceschi S, La Vecchia C. Cancer epidemiology in the elderly. Critical reviews in oncology/hematology. 2001;39(3):219–26.

33. Ukraintseva S, Yashin A. Individual aging and cancer risk: how are they related? Demographic Research. 2003;9:163–96.

34. Extermann M, Aapro M. Assessment of the older cancer patient. Hematology/oncology clinics of North America. 2000;14(1):63-77, viii-ix.

35. Cook MB, McGlynn KA, Devesa SS, Freedman ND, Anderson WF. Sex disparities in cancer mortality and survival. Cancer Epidemiol Biomarkers Prev. 2011;20(8):1629–37. doi:10.1158/1055-9965.EPI-11-0246.

36. Aizer AA, Chen M-H, McCarthy EP, Mendu ML, Koo S, Wilhite TJ et al. Marital Status and Survival in Patients With Cancer. Journal of Clinical Oncology. 2013;31(31):3869–76. doi:10.1200/JCO.2013.49.6489.

37. Kroenke CH, Michael YL, Shu X-O, Poole EM, Kwan ML, Nechuta S et al. Post-diagnosis social networks, and lifestyle and treatment factors in the After Breast Cancer Pooling Project. Psychooncology. 2017;26(4):544–52. doi:10.1002/pon.4059.

38. Nurgali K, Jagoe RT, Abalo R. Editorial: Adverse Effects of Cancer Chemotherapy: Anything New to Improve Tolerance and Reduce Sequelae? Front Pharmacol. 2018;9:245-. doi:10.3389/fphar.2018.00245.

39. Thakkar JP, McCarthy BJ, Villano JL. Age-specific cancer incidence rates increase through the oldest age groups. The American journal of the medical sciences. 2014;348(1):65–70. doi:10.1097/maj.0000000000000281.

40. Siegel R, Ma J, Zou Z, Jemal A. Cancer statistics, 2014. CA: a cancer journal for clinicians. 2014;64(1):9–29. doi:10.3322/caac.21208.

41. Qiao B, Austin AA, Schymura MJ, Browne ML. Characteristics and survival of children with acute leukemia with Down syndrome or other birth defects in New York State. Cancer epidemiology. 2018;57:68–73. doi:10.1016/j.canep.2018.10.004.

42. Xavier AC, Ge Y, Taub JW. Down syndrome and malignancies: a unique clinical relationship: a paper from the 2008 william beaumont hospital symposium on molecular pathology. J Mol Diagn. 2009;11(5):371–80. doi:10.2353/jmoldx.2009.080132.

43. Xavier AC, Taub JW. Acute leukemia in children with Down syndrome. Haematologica. 2010;95(7):1043–5. doi:10.3324/haematol.2010.024968.

44. Gamis AS. Acute myeloid leukemia and Down syndrome evolution of modern therapy--state of the art review. Pediatr Blood Cancer. 2005;44(1):13–20. doi:10.1002/pbc.20207.

45. Gormley E, Lightner D, Burgio K. American Urological Association (AUA) Guideline. Diagnosis and Treatment of Overactive Bladder (Non-Neurogenic) in Adults AUA/SUFU Guideline Linthicum, MD, USA: American Urological Association. 2014.

46. Akre O, Richiardi L. Does a testicular dysgenesis syndrome exist? Human reproduction (Oxford, England). 2009;24(9):2053–60. doi:10.1093/humrep/dep174.

47. Schnack TH, Poulsen G, Myrup C, Wohlfahrt J, Melbye M. Familial coaggregation of cryptorchidism, hypospadias, and testicular germ cell cancer: a nationwide cohort study. Journal of the National Cancer Institute. 2010;102(3):187–92. doi:10.1093/jnci/djp457.

48. Prener A, Engholm G, Jensen OM. Genital anomalies and risk for testicular cancer in Danish men. Epidemiology. 1996;7(1):14–9. doi:10.1097/00001648-199601000-00004.

49. Moller H, Prener A, Skakkebaek NE. Testicular cancer, cryptorchidism, inguinal hernia, testicular atrophy, and genital malformations: case-control studies in Denmark. Cancer causes & control : CCC. 1996;7(2):264–74.

