## Supplementary figures for "Mortality in cancer patients with congenital anomalies across different age groups: trend analysis and prognostic risk factors"

**Supplementary figure 1A, B.** Bar plots showing the number of patients died due to congenital anomalies based on site of cancer

**A)**

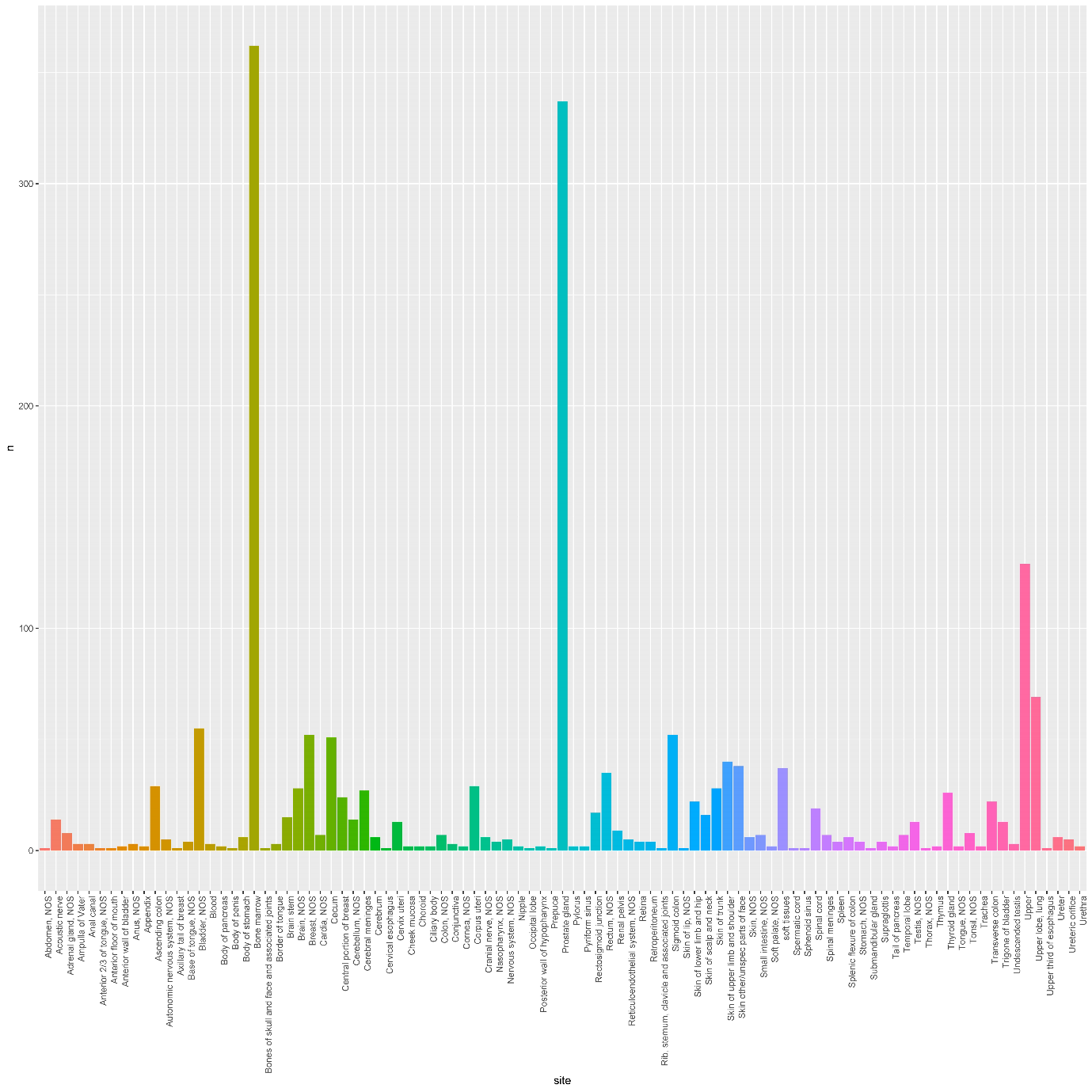

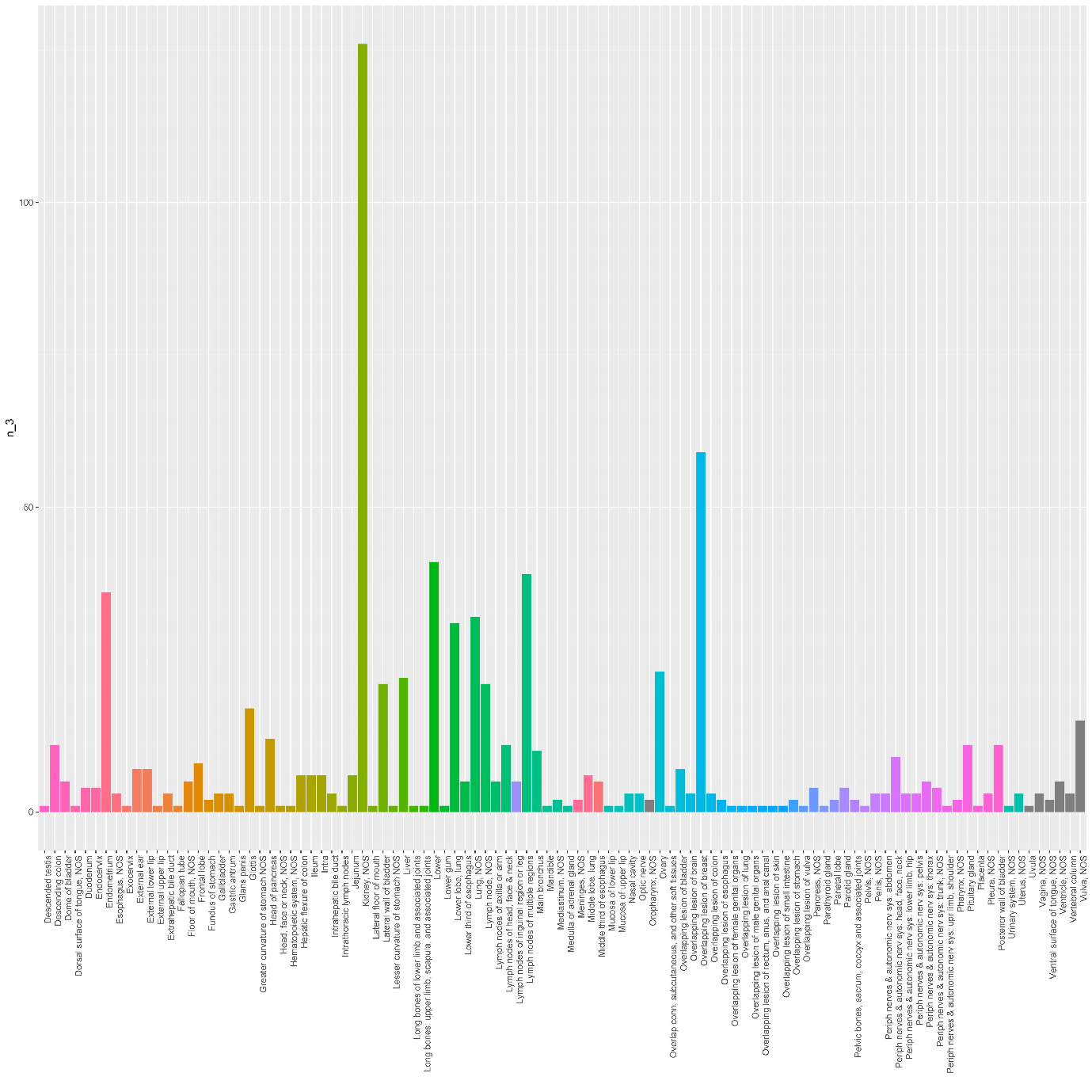
**B)**

**Supplementary figure 2.** Kaplan Meier curves comparing survival probability of congenital anomalies and other causes of death for GIT adenoma (A) and breast adenoma (B)

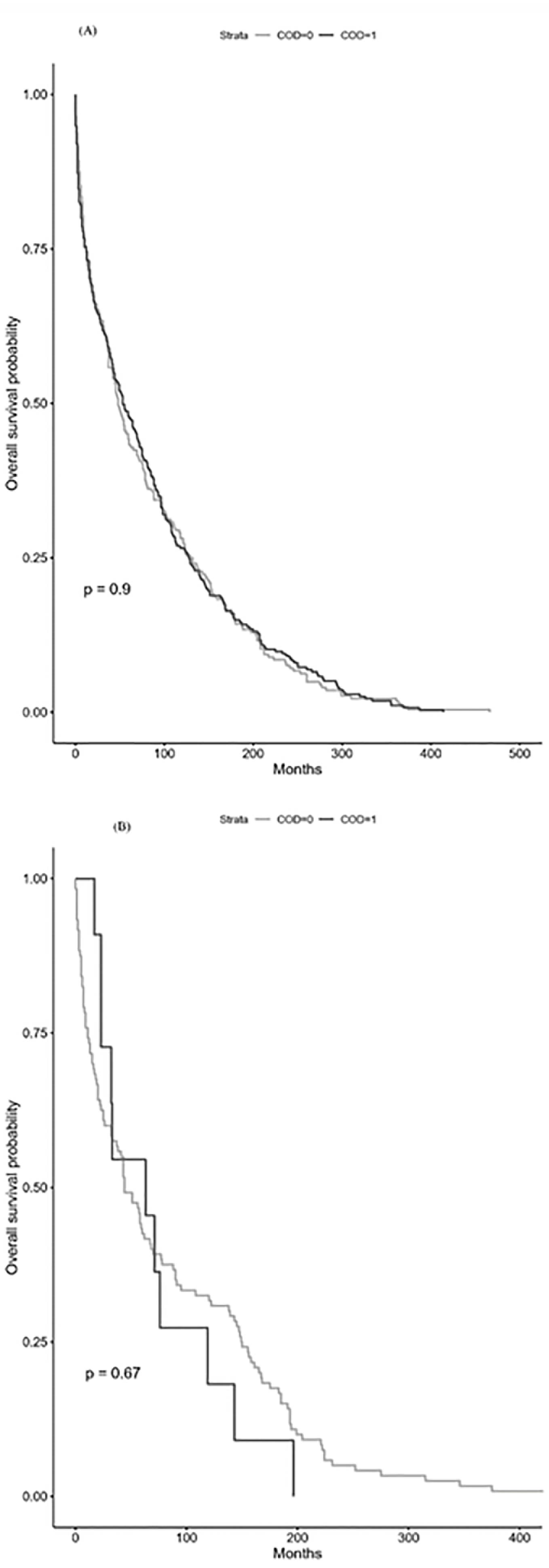
